# Longterm Temporal Dynamics of Suicidal Ideation: A Dynamic Time Warping Analysis of Depression, Anxiety, Worry, and Mastery

**DOI:** 10.64898/2026.02.20.26345909

**Authors:** M.W.M. Gijzen, A.J.C. van der Slot, M. Eikelenboom, D. de Beurs, B.W.J.H. Penninx, E.J. Giltay

## Abstract

**Background:** Suicidal ideation (SI) fluctuates over time, yet traditional static risk factors poorly align with its dynamics over time. Understanding dynamic symptom patterns may advance knowledge of the temporal interplay between SI and co-occurring symptoms in adults with depressive and anxiety disorders.

**Materials and methods:** We analyzed six waves (at baseline, and after 2, 4, 6, 9, and 13 years of follow-up) of the Netherlands Study of Depression and Anxiety (NESDA; n = 305, mean age 40.8 years, 62% female) in participants with any SI fluctuation over time. Variables included depressive, anxiety, mastery, and worry symptoms. Dynamic Time Warping (DTW) quantified within-person temporal alignment between SI and other symptoms, and an undirected network and forestplot visualized co-fluctuations. Analyses were stratified by age-groups and sex.

**Results:** Over the years, SI co-fluctuated most strongly with affective and anhedonic depressive symptoms, including sad mood, low capacity for pleasure, low general interest, pessimism, quality of mood, and decreased appetite. Select anxiety (terrified/afraid) and worry (overwhelming worries) items also aligned with SI, whereas mastery items did not. Patterns were broadly consistent across age and gender subgroups. Networks indicated that SI is part of a cluster of depressogenic symptoms but bridges to acute fear and persistent worry.

**Conclusions:** SI is a dynamic phenomenon closely linked to specific depressive, anxiety, and worry symptoms. Interventions targeting mood instability, anhedonia, and uncontrollable worry, combined with real-time monitoring, may improve personalized suicide prevention. DTW provides a framework to identify long-term temporally proximal symptom patterns.

## Introduction

Suicide is a major public health concern, with approximately 700,000 deaths annually [1]. While risk is elevated among men and individuals with prior psychiatric care, past attempts, mental health disorders, substance dependence, or socioeconomic vulnerabilities [2–10], these traditional factors remain poor predictors of suicidal thoughts and behaviours [11]. Although depression and anxiety are core predictors, with 17–20% of affected individuals reporting suicidal ideation (SI) and up to 31% in comorbid cases [12–16], static markers fail to capture the gradual emergence or intensification of SI, which fluctuates significantly on short-to-intermediate timescales [17–19].

Emotional and cognitive states, including anxiety, worry, sadness, low self-esteem, helplessness, hopelessness, and diminished control, fluctuate closely with SI [20, 21]. Despite the known importance of these proximal correlates, most evidence relies on cross-sectional assessments or two-wave designs, limiting insights into their evolution across the long-term clinical course of depression and anxiety. Transdiagnostic cognitive patterns like repetitive negative thinking (e.g. worrying and rumination) may further lower the threshold for suicidal thoughts by amplifying distress and arousal [22]. Findings remain inconsistent, particularly in MDD, despite being associated with more severe SI [22–24], persistent suicidal thoughts [25], and suicide attempts [22, 26]. While several studies report robust associations between rumination and SI independent of depression severity [26–29], others suggest that its relation to suicidal outcomes may depend on contextual factors, symptom co-occurrence, or the distinction between ideation and attempts [30, 31]. Because prior research has seldom tracked these processes over extended follow-ups, the present study contributes by monitoring within-person co-variation over years rather than identifying new risk factors. Resilience constructs like mastery, the belief in one’s ability to control life circumstances, are increasingly recognized but rarely examined longitudinally, despite evidence that higher mastery may buffer against SI severity [32–34].

While epidemiological studies such as NESDA show SI recurs frequently [16], the temporal interplay between SI, depression, anxiety, repetitive thinking, and mastery remains poorly understood. Clarifying these dynamics is crucial for identifying critical points for intervention and understanding which factors influence the onset, maintenance, or reduction of SI over time. While much recent work on temporal dynamics relies on high-frequency ecological momentary assessment (EMA), the present study uses multi-wave longitudinal data collected over thirteen years. These data capture slower, clinically meaningful shifts in depression, anxiety, worry, mastery, and SI that unfold over months or years rather than hours or days.

Traditional longitudinal models often assume synchronized timing or rely on fixed functional forms, making it difficult to compare trajectories when participants change at different rates [35].

However, long-term developmental shifts in NESDA are often unsynchronized, and individuals differ markedly in when these changes occur. Although dynamic time warping (DTW) is most commonly applied to EMA data [36–39], it provides a promising approach for multi-year designs by aligning trajectories based on their overall shape rather than fixed time points. DTW is a time-series alignment technique that enables comparison of symptom trajectories even when they evolve at different rates or times. It enables the identification of group-level temporal relationships often overlooked in traditional models [38–43]. Recent DTW applications underscore its value. DTW network analysis indicates that tightly interconnected symptoms facilitate transitions between psychiatric states [44], while intensive single-case studies identified worry and entrapment as key drivers of SI [38], supporting the Integrated Motivational-Volitional (IMV) model [45–47].

Building on these insights, the current study applies DTW to examine symptom dynamics of SI in individuals with depressive and/or anxiety disorders. We aim to identify temporal co-variation patterns between SI and depressive symptoms, anxiety, mastery, and worrying comparing these dynamic symptom networks across age and gender subgroups. Given substantial gender disparities in suicide mortality, particularly among middle-aged men, we compared four subgroups (younger/older women and younger/older men) to identify whether symptom–SI dynamics differ by gender or life stage. We hypothesize that SI co-variation patterns vary across these groups, with lower mood and mastery and higher levels of anxiety and worrying predicting stronger alignment with SI.

## Materials and methods

### Study Design and Sample

Data from the Netherlands Study of Depression and Anxiety (NESDA) were used. This is a naturalistic ongoing multisite cohort study. NESDA included a cohort of 2,981 adults (18-65 years) with a past or current diagnosis of a depressive /and or anxiety disorder (n=2,329) and healthy controls (n=652). The details of NESDA are described extensively elsewhere [48]. All participants provided written informed consent and the study was approved by the institutional ethics committee of participating universities. Baseline assessment took place from 2004-2007. Follow-up assessments were performed at 2 (wave 3; W3), 4 (W4), 6 (W5), 9 (W6) and 13 (W7) years. Participants were included in the DTW analysis if they exhibited at least one fluctuation in suicidal ideation in both SI items (i.e., based on the SSI as well as items 18 of the IDS) across the included waves within each individual. This ensured sufficient temporal variability for DTW time-series comparisons that are based on shape similarities of changes over time. Without such variability, DTW would simply cluster flat trajectories together, which is uninformative for understanding symptom dynamics. Fluctuation was operationalized at the item level. For each SI item (IDS18 and SSI item 2), we calculated the within-person standard deviation across waves. An item was considered to show fluctuation if its standard deviation was greater than zero, indicating at least one change in response category across assessments. Such changes could reflect transitions from absence to presence of SI, from presence to absence, or changes in severity, and directionality was not constrained.

### Measures

#### Suicidal thoughts

Suicidal ideation was measured in NESDA using multiple instruments that differed in both mode of administration and assessment timing: the SSI was administered by trained interviewers during structured interviews, whereas the IDS was a self-report questionnaire. As a result, the two measures were not collected on the same day. Including both instruments provided a more comprehensive assessment of SI: the SSI offered a clinician-rated evaluation of the wish to die, whereas the IDS item captured self-reported suicidal thoughts embedded within depressive symptoms, enabling convergent but non-redundant measurement.

Suicidal ideation was assessed using item 2 of the 5-item Scale for Suicide Ideation (SSI; [49]). This abbreviated version captures the presence and severity of suicidal thoughts and intentions over the past week. Item 2 assesses the wish to die (“How strong was your desire to die in the past week”). Responses are scored on a Likert scale ranging from 0 (no ideation) to higher values indicating more frequent or intense ideation. Reliability of the 5-item scale was adequate in the total sample (α ≈ .74 -.98).

Item 18 of the Inventory of Depressive Symptomatology (IDS; [50]) also measured SI. This item assesses suicidal thoughts over the past week on a 0-3 scale: 0 Does not think of suicide or death, 1 Feels life is empty or is not worth living, 2 Thinks of suicide/death several times a week for several minutes, 3 Thinks of suicide/death several times a day in depth, or has made specific plans, or attempted suicide.

#### Depression

Depressive symptoms were measured with the self-report version of the IDS. The IDS includes 29 remaining items assessing the severity of depressive symptoms experienced over the past week, such as mood, cognitive functioning, sleep disturbances, and appetite changes. Each item is rated on a 4-point scale (0–3), with higher scores indicating greater symptom severity. Internal consistency across waves was excellent (α ≈ .94 - .99).

#### Anxiety

Anxiety symptoms were assessed using the Beck Anxiety Inventory (BAI; [51]), a widely used 21-item questionnaire evaluating the severity of anxiety-related symptoms experienced during the past week. Items focus on somatic and subjective symptoms of anxiety (e.g., nervousness, dizziness, fear of losing control), rated on a 4-point scale ranging from 0 (not at all) to 3 (severely). Higher scores indicate a higher level of anxiety. Reliability of the scale was high at each wave (α ≈ .95 - .99).

#### Mastery

Perceived control over life circumstances was measured using the Pearlin Mastery Scale [52], a 7-item instrument assessing the extent to which individuals view themselves as being in control of the forces that significantly impact their lives. Items are rated on a 5-point Likert scale ranging from 1 (strongly disagree) to 5 (strongly agree), with higher scores indicating lower levels of mastery. Reliability of the scale was high at each wave (α ≈ .95 - .98).

#### Worrying

Trait worrying was assessed using the short version of the Penn State Worry Questionnaire (PSWQ-11; [53]). This short version consists only of the 11 items that assess ‘general worry’, which account for most of the variance in the PSWQ scores of the full 16-item scale [53, 54]. Each item is scored using a 0-4 Likert scale (Not at all typical of me – Certainly typical of me) with no specific recall period. Higher scores indicate higher levels of worry. Reliability of the scale was very high at each wave (α ≈ .98 - .99).

Baseline characteristics included age, sex, and education level and medication use.

### Statistical analysis

Analyses were conducted using data from six NESDA waves: baseline (W1), and follow-up waves W3, W4, W5, W6 and W7. DTW was used to analyze which individual symptoms cluster together. To examine temporal alignment between suicidal ideation (SI) and other psychological constructs, we applied Dynamic Time Warping (DTW) to participant-level time-series data. DTW is a non-linear alignment algorithm that quantifies the similarity between two temporal trajectories by finding the optimal alignment between their shapes, even when fluctuations occur at different speeds or times [37, 42, 55]. This makes DTW well suited for relatively short or unevenly spaced series, such as those available across NESDA waves.

DTW maps the relationship between symptoms in a dynamic network as it allows us to find patterns using time-series data. Within the network graphs, line thickness and color intensity represent the inverse of the mean DTW distance between symptom trajectories, such that thicker and darker edges indicate stronger temporal alignment. Only statistically significant associations are displayed. Before conducting DTW analyses, all symptom variables were scaled (z-scores) to ensure comparability and to minimize the influence of measurement scale differences. These z-scores were truncated at plus or minus 2.5 to reduce the impact of extreme values. This winsorization step reduces the impact of outliers on distance calculations and ensures more robust alignment of increases and decreases in item scores across individuals. To avoid distortions at the start and end of the series, we applied interpolation between observed assessment points. Specifically, five equally spaced interpolated values were added between each pair of observed time points before calculating DTW distances. This procedure does not introduce new information but yields smoother trajectories, reducing the potential disruptive effects of mismatched starting and endpoints and allowing DTW to more accurately capture shape similarities across time. Symptom scores were recoded where necessary to ensure consistent directionality, such that higher scores reflected greater symptom severity or distress across all constructs (i.e., higher SI, depression, anxiety, and worrying, and lower mastery).

To obtain a single SI trajectory per participant, the two standardized SI items were averaged within each wave, yielding a composite SI score. This composite trajectory was used as the sole SI outcome in all DTW analyses. DTW distances therefore reflect the temporal alignment between this composite SI signal and each individual symptom trajectory, reducing item-specific noise while preserving the temporal dynamics of SI across assessments.

DTW distances were computed between the SI time series and each of the other symptom trajectories (depression, anxiety, mastery, and worrying) within each individual. The primary outcome was the warped distance, where a smaller DTW warped distance indicates stronger temporal alignment, i.e., the two symptoms tend to rise and fall together over time. To ensure meaningful alignment and to avoid implausible overfitting, a Sakoe-Chiba band constraint of 1 time point was applied to limit the maximum allowed deviation between aligned time points, preventing overly flexible warping with scores at time points many more years apart.

In addition, linear mixed-effects models were applied to estimate the relative alignment of each symptom with the mean of the two standardized SI item scores. Models adjusted for age, sex, educational level and baseline antidepressant use, and included random intercepts for participants to account for the clustering of multiple pairwise distances within individuals. To control for spurious proximity, we also adjusted for the standard deviation of the paired item scores, given that pairs with little temporal fluctuation tend to show artificially lower distances. Results are summarized in a forest plots that presents the relative DTW distances between SI and individual symptoms in the total sample of 305 participants. To formally test whether DTW distances differed between subgroups, linear models were fitted with DTW distance as the dependent variable and subgroup membership as a categorical predictor, adjusting for within-person variability in the paired symptom trajectories. Adjusted mean DTW distances were derived from the fitted models for each subgroup, and formal pairwise comparisons between subgroups were conducted. Statistical significance (two-sided p < 0.05) identified differential temporal alignment between SI and symptoms across subgroups. Only symptoms with significantly smaller distances were interpreted as meaningfully aligned. Although our hypotheses specifically concerned smaller distances to SI (i.e., a one-sided alternative), we employed two-sided testing to account for multiple comparisons across symptoms and to adopt a conservative analytic strategy.

Participants were stratified into the following four subgroups based on gender and age at baseline:

1. Younger women (18-39 years, n = 93)
2. Middle-aged and older women (40-65 years, n = 97)
3. Younger men (18-39 years, n = 41)
4. Middle-aged and older men (40-65 years, n = 74)

All statistical analyses were performed using R version 4.3.2 (R Foundation for Statistical Computing, Vienna, Austria, 2016. URL: https://www.R-project.org/), using the packages “dtw” (version 1.23–1), “parallelDist” (version 0.2.6), “forestplot” (version 3.1.3) and “qgraph” (version 1.9.8).

## Results

### Demographic and clinical variables at baseline

The subsample included 305 participants with ≥1 fluctuation in SI. The mean age was 40.8 years (SD = 11.5), and 62.3% were women. Stratifying by sex and age revealed expected differences in mean age across subgroups.

**Table 1.** Comparison of baseline characteristics of the demographic subgroups (n=305).

| | Men < 40 | Men ≥ 40 | Women < | Women ≥ | p |
| --- | --- | --- | --- | --- | --- |
|  | N = 41 | N = 74 | 40 | 40 |  |
|  |  |  | N = 93 | N = 97 |  |
| Mean age in years (SD) | 30.6 (5.9) | 49.5 (6.4) | 29.6 (6.3) | 49.3 (6.5) |  |
| High level of education (%) | 9 (22.0) | 19 (25.7) | 36 (38.7) | 42 (43.4) | .08 |
| **Use of medication** |  |  |  |  |  |
| Antidepressants % | 31.7 | 56.8 | 39.8 | 43.3 |  |
| Benzodiazepines % | 9.8 | 16.2 | 9.7 | 15.5 |  |
| **Clinical characteristics** |  |  |  |  |  |
| Desire to die (SSI2) | .59 (.7) | .65 (.7) | .61 (.7) | .48 (.7) | .77 |
| Suicidality (IDS18) | 2.17 (.8) | 2.23 (.8) | 2.20 (.9) | 2.1 (.8) | .86 |
| Depressive symptoms (IDS) | 62.4 (12.1) | 66.0 (14.0) | 63.0 (12.7) | 63.2 (11.9) | .35 |
| Anxiety symptoms (BAI) | 39.1 (9.9) | 39.9 (9.9) | 39.5 (11.2) | 38.4 (10.0) | .81 |
| Mastery | 14.1 (3.7) | 13.8 (3.7) | 14.2 (4.0) | 14.2 (4.5) | .95 |
| Worrying (PSWQ) | 41.1 (10.1) | 38.5 (9.1) | 40.3 (9.2) | 35.9 (9.3) | **.004** |

*Note.* For continuous outcomes (worry, mastery, depressive symptoms, and anxiety), one-way analyses of variance (ANOVA) were conducted, followed by Tukey post-hoc comparisons. Pearson's chi-Square test was used for categorical variables (desire to die, suicidality, and level of education). SSI: Scale for Suicidal Ideation; IDS: Inventory of Depressive Symptomatology; BAI: Beck Anxiety Inventory; PSWQ: Penn State Worry Questionnaire. Ranges: IDS (27-120), BAI (21-84), PSWQ (11-55) Pearlman Mastery Scale (5-25).

### Overall symptom network structure

To gain an overview of cross-symptom dynamics, we aggregated all individual DTW networks into a single, undirected graph that retained only edges with significantly shorter DTW distances than expected by chance. The resulting network (Figure 1) shows that most symptoms clustered within their original domains (e.g., IDS for depression, BAI for anxiety, Mastery, and ‘W’ for worry), but important cross-domain connections were also present. The SI-items (IDS18 and SSI2), highlighted in red, were connected to each other and to several key depressive symptoms. These included core depressive symptoms such as *sad mood* (IDS5), *quality of mood* (IDS10), *low capacity for pleasure* (IDS21), *responsiveness of mood* (IDS8), and *low general interest* (IDS19). The mastery item *can’t change my life* (Mastery3) and the worry item *overwhelming worries* (W1) were connected to a *desire to die* (SSI2). Anxiety links with SI were more selective, involving primarily cognitive-affective items such as *terrified/afraid* (BAI9), *feeling scared* (IDS17), *fear of losing control* (BAI14) and *faint/lightheaded* (BAI19) and arousal items such as *shaky, unsteady* (BAI13)*, feeling of choking* (BAI11), *hands trembling* (BAI12) and *numbness or tingling* (BAI1).

**Figure 1.**
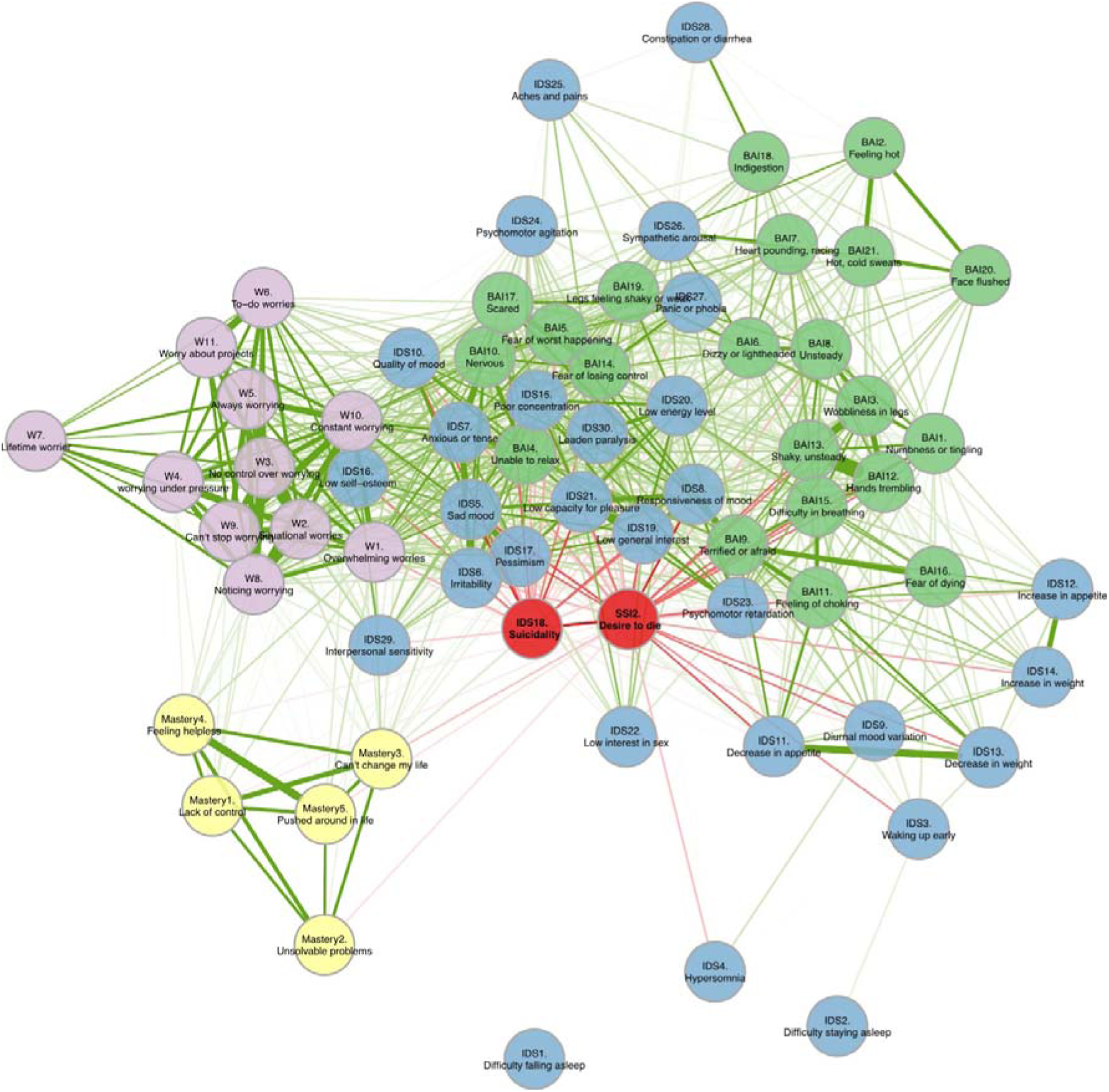
Undirected network of depressive symptoms, anxiety symptoms, worry and mastery with suicidal ideation. Undirected network based on pooled data of all participants (n=305) of suicidal thoughts (IDS18), desire to die (SSI2), anxiety (BAI), depression (IDS), mastery, and worry (PSWQ). Each node represents a single item. The thickness of edges represent the strength of the co-variation pattern over time (i.e., the inverse of the mean distance) and were color-coded: red edges indicate edges directly connected to the two SI items. All edges represent positive co-alignment between item scores over time. Node color separates symptom domains.

### Symptom-specific alignment with SI

We next examined which individual symptoms showed the strongest temporal correspondence with SI by analyzing forest plots of DTW distances adjusted for age, sex, and educational level (Figure 2). In this framework, smaller distances reflect closer symptom-SI coupling over time.

**Figure 2.**
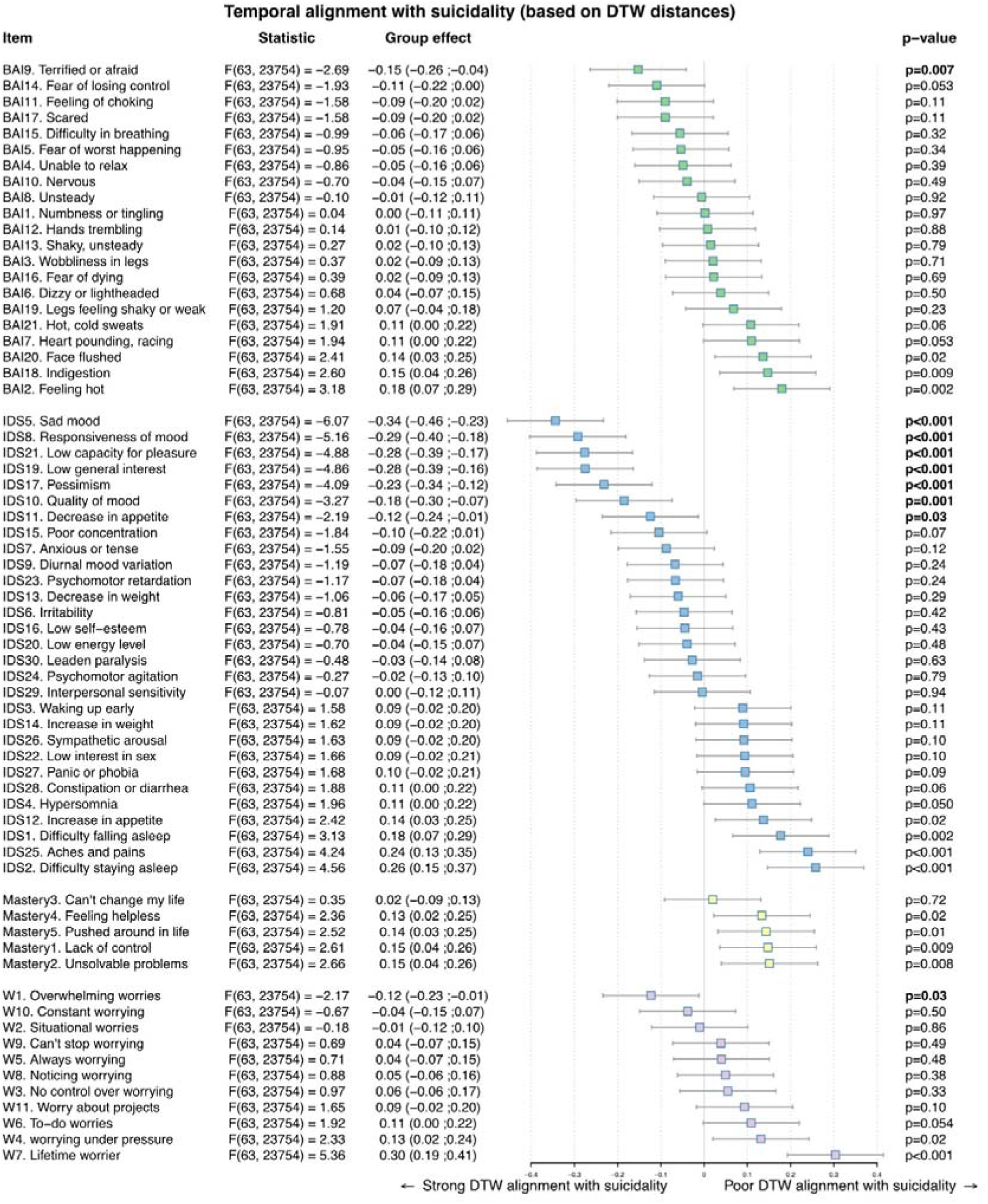
Overview of dynamic distances from suicidal ideation for all items across domains. This figure summarizes the dynamic time warping (DTW) distances between individual items and suicidal ideation (SI) across all measured symptom domains: anxiety (BAI), depression (IDS), mastery, and worry (PSWQ). Colors separate symptom domains. Smaller distances indicate greater temporal alignment (co-fluctuation) with SI. Error bars represent 95% confidence intervals around the estimated DTW distances. Group effect statistics are shown as F-values from ANOVA tests. Corresponding p-values are indicated; p < 0.05 was considered statistically significant, which are interpreted as meaningfully covarying over time with SI.

Significant co-fluctuations with SI over time were observed for affective depression symptoms, with *Sad mood* (IDS5; *p* < .001), *Responsiveness of mood* (IDS8; p < .001), *Low capacity for pleasure* (IDS21; *p* < .001), *Low general interest* (IDS19; *p* < .001), *Pessimism* (IDS17; *p* = .001), *Quality of mood* (IDS10; *p* = .001), and *Decrease in appetite* (IDS11; *p* = .03). Beyond depressive symptoms, anxiety symptom *Terrified or afraid* (BAI9; *p* = .007) and worry symptoms *Overwhelming worries* (W1; *p* = .03) emerged as significantly aligned with SI. In contrast, none of the mastery items showed significant alignment to SI.

### Subgroup analyses: age and sex

When subgroup differences were examined, the same set of symptoms namely depressive symptoms (*Sad mood, Responsiveness of mood, Low capacity for pleasure, Low general interest, Pessimism, Quality of mood, and Decrease in appetite*), anxiety (*Terrified or afraid*), and worrying (*Overwhelming worries*) consistently showed significant alignment with SI in all four subgroups. None of the mastery items demonstrated such proximity (all p > .05).

Exploratory post-hoc tests indicated that the strength of these associations was broadly similar across men and women and across younger and older participants. Younger women showed more proximity to SI than older women and older men for the depressive symptom *Responsiveness of mood* (IDS8) and also more proximity to SI than young men for low capacity for pleasure (see Supplementary Figures 1–4 for details).

## Discussion

This study aimed to identify specific symptom dynamics of SI by examining within-person co-fluctuations over several years. Findings emphasize that suicidality does not merely track overall depression severity but fluctuates in tandem with specific symptoms. The strongest temporal alignments were observed with affective depressive symptoms, including *sad mood*, *reduced responsiveness of mood*, *low capacity for pleasure*, *low general interest*, *pessimism*, *quality of mood*, and *decrease in appetite*. These results reinforce the view that suicidality is more closely tied to persistent negative affect and anhedonia than other features. Additionally, the anxiety symptom *terrified or afraid* and the cognitive worry item *overwhelming worries* showed significant alignment with SI, suggesting that acute fear responses and transdiagnostic cognitive patterns play complementary roles in the dynamics of suicidality.

Consistent with prior DTW research, we found that sad mood, low interest, and overwhelming worry co-fluctuated with SI also over shorter time intervals [56]. While core affective pathways appear robust, additional symptom–SI associations may depend on sample characteristics and symptom variability. For instance, our observed link between decreased appetite and SI aligns with earlier findings [44], though direct comparisons are limited when studies include participants without meaningful symptom fluctuations [44].

Reduced mood responsiveness, reflecting blunted reactions to positive events, co-fluctuated with SI alongside other core depressive symptoms. Network studies consistently identify sad mood and anhedonia as central to depression [57–65] and anhedonia is strongly linked with suicidal thoughts in clinical and observational studies [66, 67]. Notably, recent changes (past two weeks) in anhedonia, rather than stable levels, best predict SI [68], while positive affect can trigger broader network-wide improvements [63, 69, 70]. Although non-core depressive symptoms often improve after core symptoms [71], SI improvements may precede core changes or occur independently of overall depression trajectories [72]. Together, these findings underscore the need to move beyond static sum scores and focus on the dynamic interplay between symptoms and SI within individuals.

Beyond affective symptoms, *terrified or afraid* (BAI9) and *overwhelming worries* aligned closely with SI, suggesting acute fear responses and persistent worry represent complementary cognitive–affective pathways to suicidal thoughts. Network analyses similarly identify sad mood and worry as central symptoms in depression and anxiety in psychiatric samples [73], with uncontrollable worry as a key bridging symptom [74]. Furthermore, worry controllability is a strong transdiagnostic predictor of SI [75]. These converging findings support our observation that overwhelming worries co-fluctuated with SI and highlight the importance of targeting cognitive patterns related to worry.

By contrast, mastery showed no temporal alignment with SI, likely reflecting differences in timescales. Mastery is conceptualized as a relatively stable trait [52], whereas SI fluctuates rapidly, sometimes across hours or days [19, 76]. While low mastery contributes to increased risk for suicidal thoughts [33], it may influence vulnerability indirectly rather than co-fluctuating with SI in the moment. This mismatch highlights a broader methodological point: constructs operating on different temporal scales may not show proximal associations in dynamic analyses [77, 78]. Mastery may also operate indirectly by influencing more proximal emotional states (e.g., anxiety, negative affect). In the symptom network (Figure 1), mastery items was especially linked to worry and depressive items, which subsequently showed links to SI. As such, mastery’s association with SI may be mediated by these nearer-term affective processes, which could explain why mastery did not show strong proximal alignment in our DTW analyses.

In this study, we explored how symptoms fluctuate and relate to one another over years using DTW. Unlike previous research relying on cross-sectional or limited longitudinal designs (e.g. 2-3 assessments) that capture group averages, our DTW analysis quantifies individual temporal alignment before group aggregation. This “within-person first” approach is essential due to the ergodicity problem, as group-level associations often fail to reflect individual processes over time [79, 80].

Furthermore, because symptom fluctuations are rarely synchronous, averaging data beforehand obscures critical individual dynamics. Unlike standard time-series methods, DTW handles the uneven time intervals common in psychiatric research [55]. Building on EMA evidence of heterogenous depressive symptom behaviour over short periods [81], our findings demonstrate that DTW reveals meaningful co-fluctuating patterns even in datasets collected less frequently over longer periods.

Several limitations warrant consideration. First, while DTW accommodates unevenly spaced data, the number and spacing of assessments may have limited sensitivity to subtle temporal effects. Second, the analyses remain correlational, precluding causal inference between symptoms and SI. Third, the lack of proximal alignment for mastery should be interpreted cautiously, as measurement timing, item phrasing, or statistical power may contribute. Finally, the subgroup of our cohort which oversampled subjects with depression and anxiety may limit generalizability to community populations.

To conclude, our findings highlight that SI most strongly co-fluctuates over years with specific affective, anhedonic and cognitive worry symptoms. These results underscore the value of considering both symptom-specific and temporal dynamics when assessing suicidal thoughts and designing interventions. Clinically, interventions targeting mood instability, anhedonia, and uncontrollable worry may be especially effective in reducing SI. Integrating such approaches with frequent or real-time monitoring could further support personalized suicide prevention [38, 76, 82].

## Data availability

The data that support the findings of this study are available from the NESDA consortium (www.nesda.nl), but restrictions apply to the availability of these data, which were used with specific permission for the current study and are not publicly available. Due to privacy restrictions and ethical regulations, the raw data are not publicly available. Access to the data can be requested through the respective study secretariats, subject to approval by their scientific committees and in accordance with their data access protocols.

## Acknowledgements

Funding and acquisition of the ZonMw Grant that supported this study was done by Prof. Dr. Diana van Bergen (University of Groningen).

