## Supplemental materials for "Longterm Temporal Dynamics of Suicidal Ideation: A Dynamic Time Warping Analysis of Depression, Anxiety, Worry, and Mastery"

**Figure S1.** Dynamic alignment of depressive symptoms with suicidal ideation across sex and age groups.

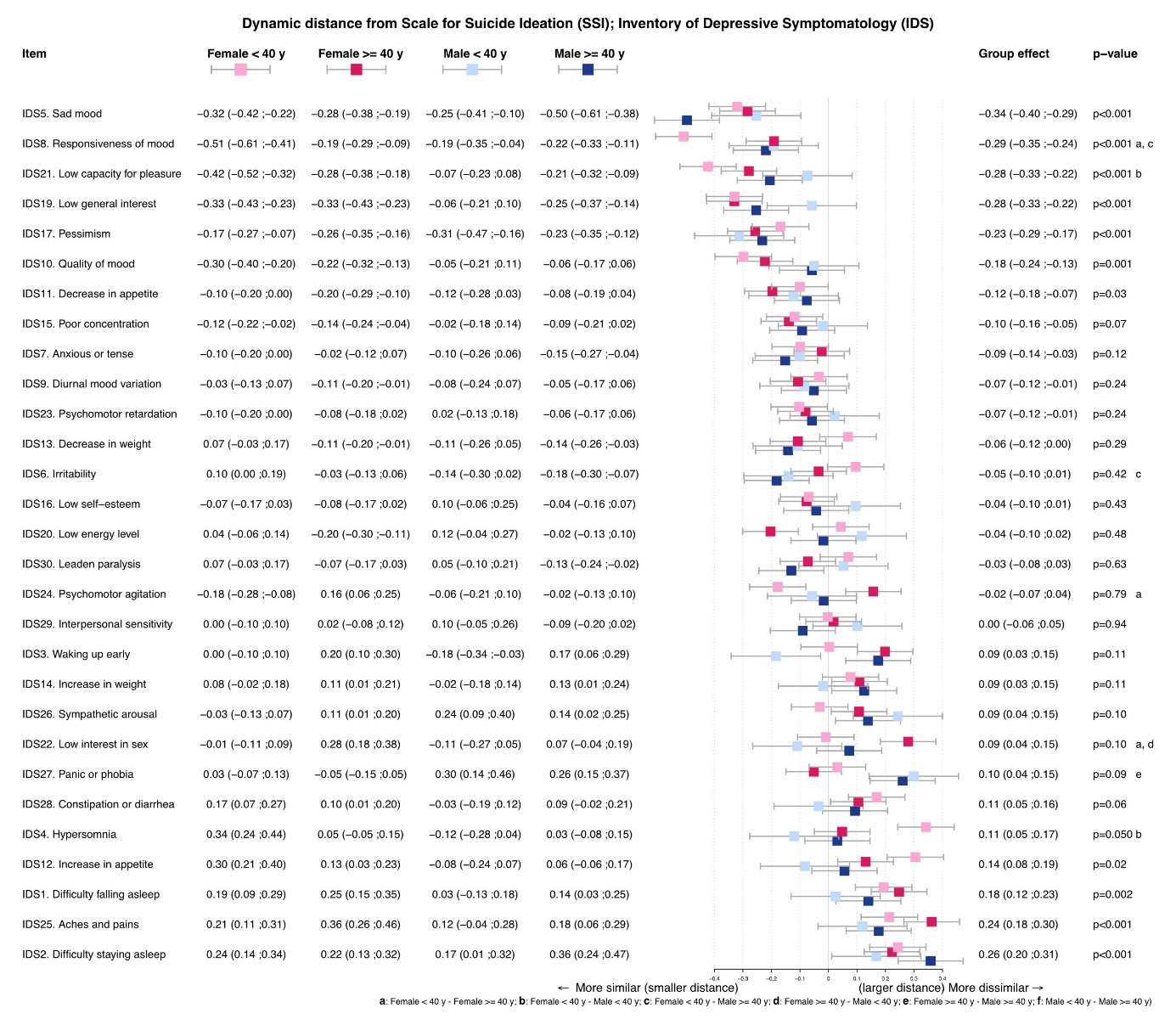

Undirected DTW analysis was used to compute the temporal distance between depressive symptoms (IDS items) and suicidal ideation (SI). Group effects by sex (male, female) and age (<40 years, 40-65 years) are indicated. Smaller distances indicate more similar fluctuations within individuals over time. P-values for the item compared at the group-level with SI are displayed to the right of each item, with p < 0.05 considered statistically significant compared to all other items. Sub-group effects are provided and statistically significant post-hoc comparisons are indicated with a letter on the right of the overall p value of each item. The symptoms, “sad mood” (IDS5, p < .001), “responsiveness of mood” (IDS8, p < .001),”low capacity for pleasure” (IDS9, p < .001), “low general interest” (IDS19, p < .001), “pessimism” (IDS17, p = .01), “quality of mood” (IDS10, p = .001) “decrease in appetite” (IDS11, p = .03) were significantly aligned with SI across groups. Significant subgroup differences were found for “responsiveness of mood” (IDS8),”low capacity for pleasure” (IDS9).
a = between younger women (40-) and older women (40-65), b = between younger women (40-) and younger men (40-), c = between younger women (40-) and older men (65+) d = between older women (40-65) and younger men (40-), e = between older women (40-65) and older men (40-65), and f = younger men (40-) and older men (40-65).

**Figure S2.** Dynamic alignment of anxiety symptoms with suicidal ideation across sex and age groups.

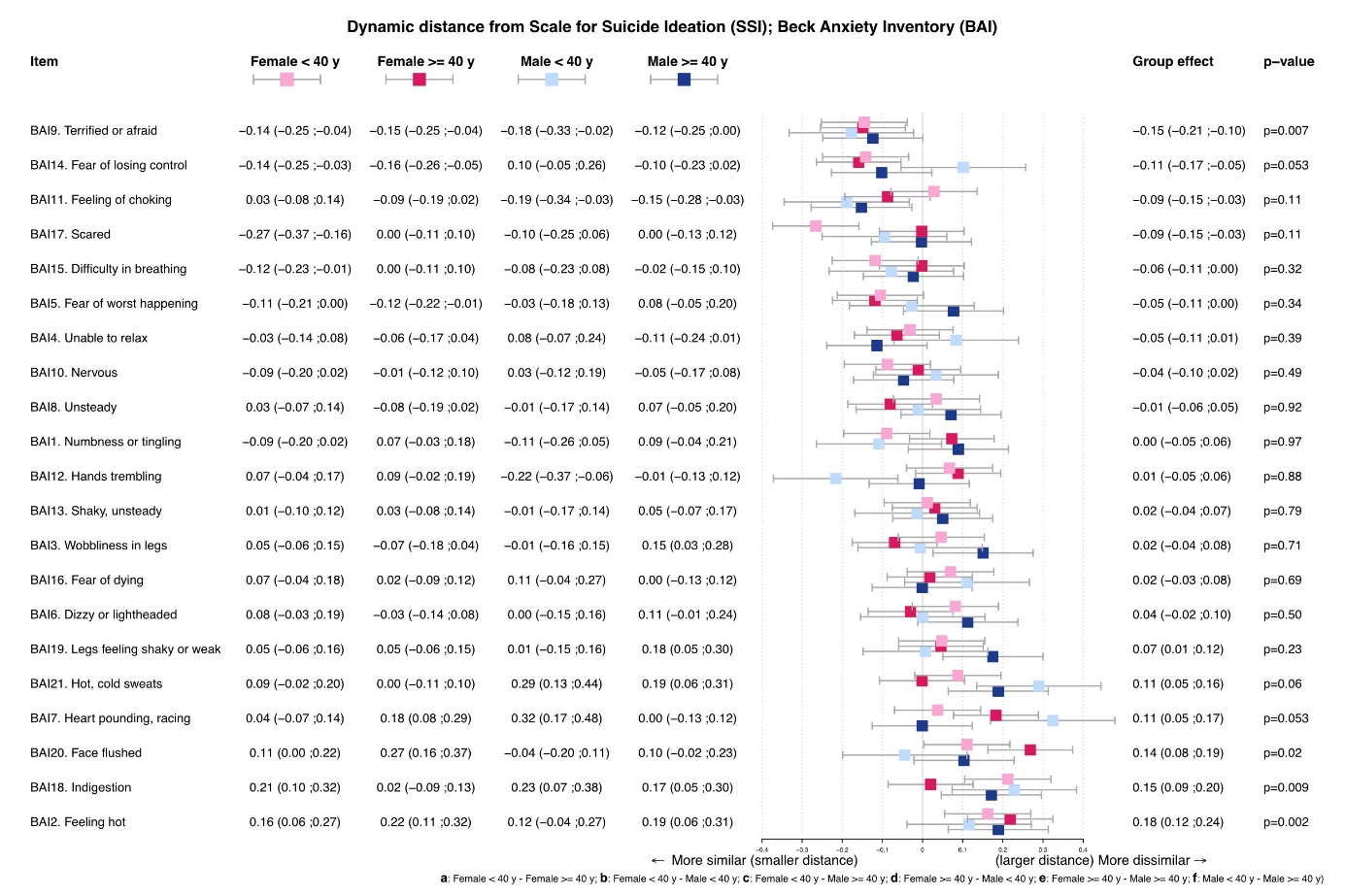

Undirected DTW analysis was used to compute the temporal distance between anxiety symptoms (BAI items) and suicidal ideation (SI). Group effects by sex (male, female) and age (<40 years, 40-65 years) are indicated. Smaller distances indicate more similar fluctuations within individuals over time. P-values for the item compared at the group-level with SI are displayed to the right of each item, with p < 0.05 considered statistically significant compared to all other items. Sub-group effects are provided and statistically significant post-hoc comparisons are indicated with a letter on the right of the overall p value of each item. The symptom of “feeling terrified or afraid” (BAI9, p = .007) was significantly aligned with SI. No significant subgroup differences were found.
a = between younger women (40-) and older women (40-65), b = between younger women (40-) and younger men (40-), c = between younger women (40-) and older men (65+) d = between older women (40-65) and younger men (40-), and e = between older women (40-65) and older men (40-65).

**Figure S3.** Dynamic alignment of mastery with suicidal ideation across sex and age groups.

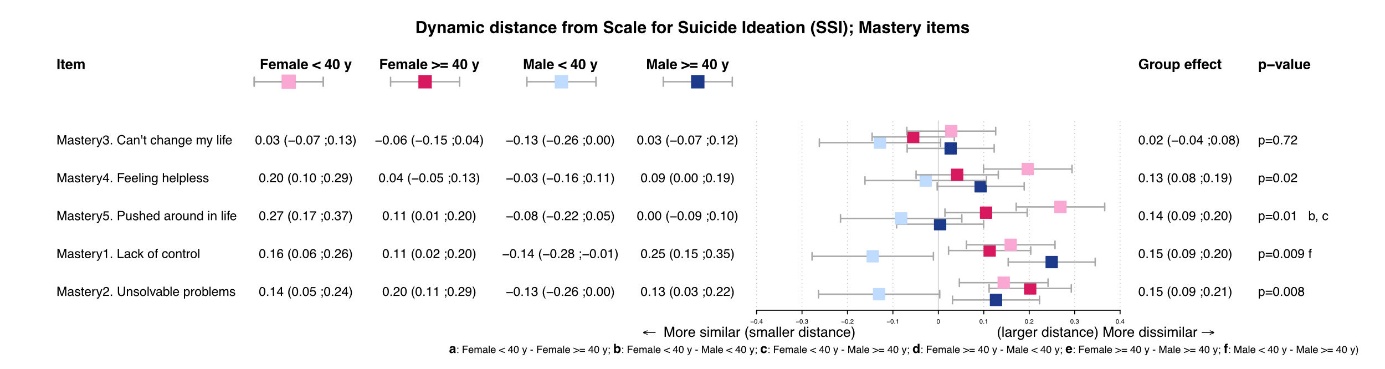

Undirected DTW analysis was used to compute the temporal distance between mastery (Pearlin Mastery Scale items) and suicidal ideation (SI). Group effects by sex (male, female) and age (<40 years, 40-65 years) are indicated. Smaller distances indicate more similar fluctuations within individuals over time. P-values for the item compared at the group-level with SI are displayed to the right of each item, with p < 0.05 considered statistically significant compared to all other items. Sub-group effects are provided and statistically significant post-hoc comparisons are indicated with a letter on the right of the overall p value of each item. No mastery symptoms were significantly aligned with SI (p > .001). No significant subgroup differences were found.

a = between younger women (40-) and older women (40-65), b = between younger women (40-) and younger men (40-), c = between younger women (40-) and older men (65+).

**Figure S4.** Dynamic alignment of worry symptoms with suicidal ideation across sex and age groups.

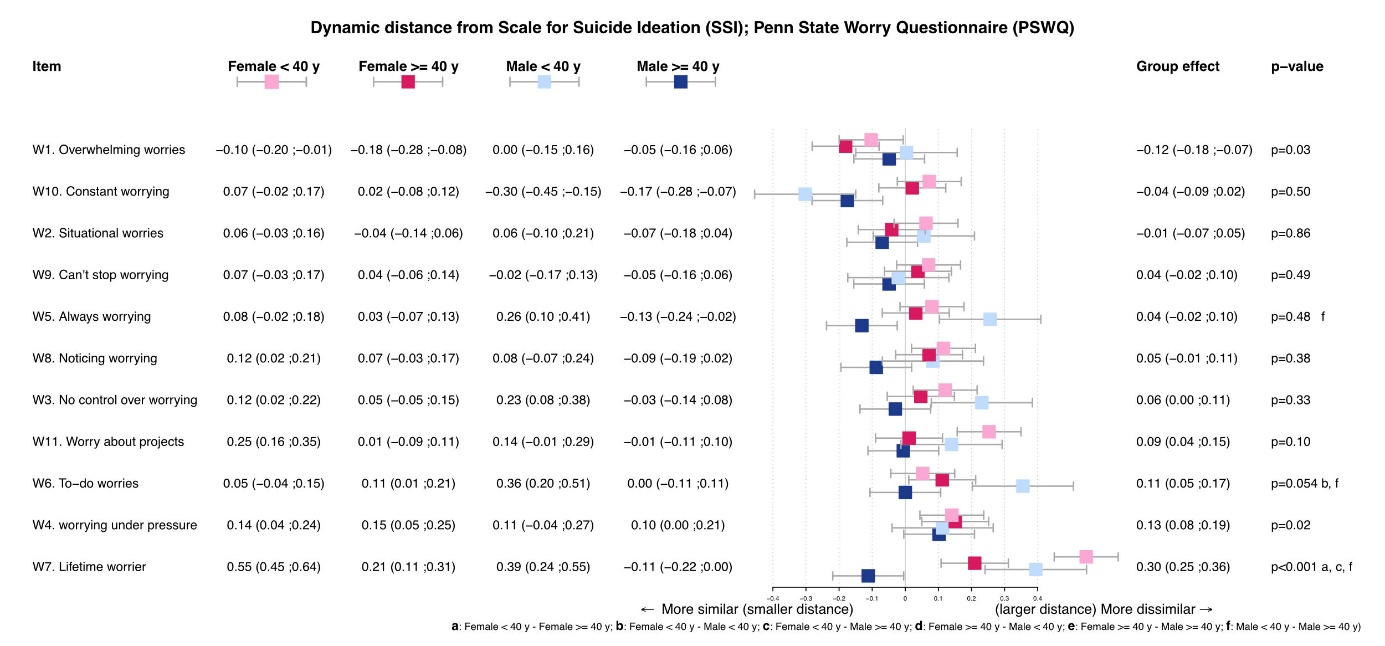

Undirected dynamic time warping (DTW) analysis was used to compute the temporal distance between individual items of the Penn State Worry Questionnaire (PSWQ) and suicidal ideation (SI). Group effects by sex (male, female) and age (<40 years, 40-65 years) are indicated. Smaller distances indicate more similar fluctuations within individuals over time. P-values for the item compared at the group-level with SI are displayed to the right of each item, with p < 0.05 considered statistically significant compared to all other items. Sub-group effects are provided and statistically significant post-hoc comparisons are indicated with a letter on the right of the overall p value of each item. The symptom of “overwhelming worries” (W1, p = .03) was significantly aligned with SI. No significant subgroup differences were found.

a = between younger women (40-) and older women (40-65), b = between younger women (40-) and younger men (40-), c = between younger women (40-) and older men (65+) d = between older women (40-65) and younger men (40-), e = between older women (40-65) and older men (40-65), and f = younger men (40-) and older men (40-65).
